# Racial Disparities and Trends in Anticoagulant Use among Ambulatory Care Patients with Atrial Fibrillation and Atrial Flutter in the United States from 2007-2019

**DOI:** 10.1101/2024.06.14.24308960

**Authors:** Vincent Kan, Kate Lapane, David McManus, Jonggyu Baek, Chad Darling, Matthew Alcusky

## Abstract

**Introduction:** Atrial fibrillation (AF) is the most common sustained cardiac arrhythmia, significantly increasing the risk of stroke. The introduction of direct oral anticoagulants (DOACs) since 2010 has transformed anticoagulation therapy, offering an alternative to warfarin with improved safety profiles. Despite the increased adoption of DOACs, disparities in their use among different racial and ethnic groups in the United States remain understudied.

**Methods:** This study utilized a repeated cross-sectional design, analyzing data from the National Ambulatory Medical Care Survey (NAMCS) from 2007 to 2019. The study population included adults diagnosed with AF or atrial flutter (AFL). We analyzed the temporal trends of DOAC and warfarin use from 2007 to 2019. We examined the prevalence of DOAC versus warfarin use and assessed associations between race/ethnicity, patient characteristics, and DOAC utilization from 2011 to 2019. Multivariable modified Poisson regression models were used to calculate adjusted prevalence ratios (aPR) for the associations.

**Results:** From 2011 to 2019, NAMCS recorded 3,224 visits involving AF or AFL, representing a weighted estimate of 103.6 million visits. DOAC use increased significantly, with apixaban becoming the predominant anticoagulant by 2016. Non-Hispanic Black patients were less likely to use DOACs compared to non-Hispanic White patients over time (aPR 0.75; 95% CI, 0.63-0.90). Patients with Medicaid insurance were also less likely to use DOACs (aPR 0.14; 95% CI: 0.04-0.46).

**Conclusion:** Despite the shift from warfarin to DOACs for AF and AFL treatment, significant racial and socioeconomic disparities persist. Non-Hispanic Black patients and those with Medicaid insurance are less likely to use DOACs. These findings highlight the need for targeted strategies to ensure equitable access to advanced anticoagulant therapies.

## INTRODUCTION

Atrial fibrillation (AF) is the most common sustained cardiac arrhythmia, with a lifetime risk of 1-in-4 for those over 40 years of age.^1^ If left untreated, AF patients face a five-fold increased risk of stroke compared to those without AF. Strokes related to AF are also devastating, resulting in higher morbidity and mortality rates than strokes from other causes.^2,3^ Anticoagulation is indicated for patients with AF with two or more stroke risk factors, as identified by a CHA_2_DS_2_-VASc risk score. The landscape of chronic oral anticoagulation has changed significantly since the FDA approved of the first direct oral anticoagulants (DOACs) in 2010. Dabigatran and rivaroxaban are non-inferior to warfarin for stroke prevention and systemic embolism and have equivalent or superior rates of adverse events compared to warfarin.^4,5^ Apixaban has been demonstrated to outperform warfarin for stroke prevention and is associated with lower rates of bleeding.^6^ Over the last fourteen years, and owing to their favorable safety profile, fewer drug-drug interactions, and less complex monitoring regimen, DOACs have become the predominant form of oral anticoagulation for non-valvular AF.

Previous studies have shown increasing use of oral anticoagulants overall among patients with AF, with increasing adoption of DOACs and a gradual decrease in the use of warfarin after the approval of dabigatran. Furthermore, racial and ethnic disparities in the initiation of DOAC among AF patients have been shown in clinical trial registries and the Veterans Health Administration (VA) System,^7,8^ and among those who have been diagnosed with venous thromboembolism.^9^ However, data on oral anticoagulant use among the general population among different racial and ethnic groups in the United States beyond 2014 are limited, particularly regarding the use of apixaban, which was FDA-approved in December 2012.^10–16^

Using a nationally representative ambulatory care database, this study aims to examine changes in anticoagulant use from 2007 to 2019 among patients with AF and atrial flutter (AFL). Specifically, the study will explore the prevalence of DOAC versus warfarin use and quantify associations between race/ethnicity and other patient characteristics and DOAC use.

## METHODS

### Study Design and Data Source

This is a repeated cross-sectional study using data from a nationally representative survey, the National Ambulatory Medical Care Survey (NAMCS), from 2007-2019 (excluding 2017 due to data being unavailable) to summarize and analyze recent time trends in the usage of DOACs for AF and AFL. NAMCS is an annual cross-sectional survey of non-federally employed, office-based physician visits conducted by the National Center for Health Statistics of the Centers for Disease Control and Prevention (CDC). As of 2019, NAMCS classifies physicians into fourteen specialty groups: general and family practice, internal medicine, pediatrics, general surgery, obstetrics and gynecology, orthopedic surgery, cardiovascular diseases, dermatology, urology, psychiatry, neurology, ophthalmology, otolaryngology, and all other. NAMCS utilizes a multistage probability design in which a sample of patient visits are randomly selected with a pre-specified probability from within certain physician practices, which in turn are chosen randomly with pre-specified probability from within primary sampling units across the country.^17,18^ Data are collected on patients and visits, and physician practice characteristics.

### Study Population

We included adults aged 18 years and older with a listed diagnosis of AF or AFL. NAMCS collects up to three diagnoses for each visit using International Classification of Diseases, Ninth Revision, Clinical Modification (ICD-9-CM) codes from 2006-2013, up to five diagnoses using ICD-9-CM codes from 2014-2015, and up to five diagnoses using ICD-10-CM codes from 2016-2019. AF and ALF were defined using ICD-9-CM codes 427.31 and 427.32 and ICD-10-CM codes 148.0, 148.1, 148.2, 148.3, 148.4, 148.91, 148.92. To make our findings comparable across the years, our primary analysis included the first three diagnoses listed for each visit, an approach similar to prior studies utilizing NAMCS.^19–21^ Given the change in the number of diagnoses recorded by NAMCS over the years in our study, we conducted a sensitivity analysis to include the maximum number of diagnoses collected in the NAMCS each year.

### Anticoagulant use

Our primary outcome is the overall use of oral anticoagulants by type. We classified patients as users or non-users of oral anticoagulants, including apixaban, dabigatran, rivaroxaban, edoxaban, and warfarin, based on whether any of these medications were recorded on their NAMCS visit records. NAMCS collected up to eight medications prescribed, ordered, or continued in the 2006-2011 surveys, up to ten medications in the 2012-2013 surveys, and up to thirty medications in the 2014-2019 surveys. To make our findings comparable across the years, our primary analysis included the first eight medications listed for each visit, an approach similar to prior studies utilizing NAMCS.^19–21^ Given the change in the number of medications recorded by NAMCS over the years in our study, we conducted a sensitivity analysis to include the maximum number of medications collected in the NAMCS each year.

### Baseline characteristics

Patient demographic characteristics included age (18-64, 65-74, ≥75 years), sex (female, male), antiplatelet use (aspirin, clopidogrel, prasugrel, ticagrelor, aspirin/dipyridamole), race/ethnicity ( Hispanic, Non-Hispanic Black, Non-Hispanic White, Other including Asian, Native Hawaiian or Other Pacific Islander, American Indian or Alaska Native, more than one race), medical comorbidities (chronic kidney disease/end-stage renal disease, congestive heart failure, chronic obstructive pulmonary disease, prior cerebral vascular disease or transient ischemic attack, diabetes, hypertension), payment source (Medicare, Medicaid, private insurance, self-pay, and other), and anticoagulant use (warfarin, DOAC, not treated). We did not include region or physician specialty data due to a lack of availability for 2018 and 2019 in NAMCS.

### Statistical Analysis

We analyzed the use of anticoagulants and baseline characteristics across different racial and ethnic groups. Continuous variables were presented as means ± standard deviation and categorical variables as percentages. To examine temporal trends in the use of oral anticoagulants – including warfarin and DOACs – among patients with AF and AFL patients, we plotted the prevalence estimates from 2007 to 2019. We used a modified Poisson regression model to calculate adjusted prevalence ratios for the associations between race/ethnicity, age, sex, primary payor, medical comorbidities, and year with DOAC use from 2011 to 2019, beginning after the introduction of the first DOAC in late 2010. We also included a year and race/ethnicity interaction term in the model to examine the temporal variation between race/ethnicity and DOAC use. A type III test was used to determine the significance of the interaction term. Data were weighted to obtain nationally representative annual results using survey procedures (*svy* commands) in Stata/MP Version 17 (StataCorp, College Station, TX). In our analysis, we combined multi-year survey data, accounting for variations in sample sizes and design by utilizing the weighted average approach. Each year’s data was weighted according to its specific sampling weights, ensuring that the contribution of each dataset was proportional to its representative value in the overall population.

## RESULTS

### Sample Characteristics

Between 2011 and 2019, the NAMCS included data on 267,485 visits to office-based physicians. Of those, 3,224 visits involved patients who had an AF or AFL diagnosis, which represents a total weighted estimate of 103.6 million visits. Between 2011 and 2019, more men had outpatient visits for AF or AFL than women (Table 1). An overall average of 45% of those visits had an anticoagulant prescription recorded. The mean age of patients was 73.3. About two-thirds (70%) of the visits were covered by Medicare. Non-Hispanic Whites, Non-Hispanic Blacks, and Hispanics accounted for 84.5%, 6.5%, and 5.9% of the visits, respectively.

**Table 1.** Comparison of Baseline Characteristics by Race/Ethnicity for Patients with Atrial Fibrillation and Atrial Flutter, 2011 - 2019.

|  | Overall | White | Black | Hispanic | AANHPI/<br>AIAN/Multi<sup>1</sup> |
| --- | --- | --- | --- | --- | --- |
| Unweighted, n (%) | 3,224 | 2,828 (87.7) | 172 (5.3) | 141 (4.3) | 82 (2.6) |
| Weighted, n (%) | 103,617,866 | 87,508,793<br>(84.5) | 6,773,908<br>(6.5) | 6,076,065<br>(5.9) | 3,259,100<br>(3.2) |
| Age, years, mean (SE) | 73.6 (±11.7) | 73.9 (±11.5) | 70.4 (±12.6) | 71.4 (±14.1) | 70.6 (±11.8) |
| 18-65, % | 20.5 | 19.9 | 17.0 | 23.7 | 38.8 |
| 65-74, % | 29.0 | 28.8 | 31.1 | 27.9 | 32.1 |
| >75, % | 50.5 | 51.4 | 52.0 | 48.5 | 29.1 |
| Sex, % |  |  |  |  |  |
| Female | 43.5 | 42.6 | 49.8 | 51.5 | 40.5 |
| Male | 56.6 | 57.4 | 50.2 | 48.5 | 59.5 |
| Antiplatelet use, % |  |  |  |  |  |
| Aspirin | 17.8 | 18.5 | 15.3 | 16.4 | 15.7 |
| Other | 3.1 | 2.8 | 4.2 | 5.9 | 3.2 |
| Medical comorbidities<sup>2</sup>,<br>% |  |  |  |  |  |
| CKD/ESRD | 11.4 | 9.4 | 25.8 | 20.2 | 19.7 |
| CHF | 21.8 | 21.4 | 22.9 | 22.9 | 29.3 |
| COPD | 12.2 | 13.1 | 6.8 | 8.5 | 8.0 |
| CVA/TIA | 8.8 | 7.7 | 8.1 | 23.5 | 13.3 |
| Diabetes | 74.8 | 73.5 | 85.4 | 81.0 | 77.2 |
| Hypertension | 71.1 | 69.7 | 82.9 | 80.0 | 66.7 |
| Reason for visits, % |  |  |  |  |  |
| Acute problem | 12.3 | 11.0 | 11.1 | 25.7 | 20.0 |
| Chronic problem | 68.5 | 70.7 | 69.7 | 52.9 | 56.3 |
| Other | 19.3 | 18.2 | 29.2 | 21.4 | 23.7 |
| Insurance coverage, % |  |  |  |  |  |
| Medicaid | 1.8 | 1.2 | 3.0 | 5.0 | 9.5 |
| Medicare | 70.0 | 71.4 | 67.6 | 69.0 | 38.8 |
| Private | 21.1 | 20.8 | 16.3 | 22.3 | 36.7 |
| Self-pay/Other | 7.2 | 6.6 | 13.1 | 3.7 | 15.0 |
| Anticoagulant use, % |  |  |  |  |  |
| Warfarin | 25.9 | 25.6 | 36.9 | 23.3 | 16.5 |
| DOAC | 18.9 | 19.6 | 4.6 | 21.0 | 21.8 |
| Not treated | 55.2 | 54.7 | 58.6 | 55.7 | 61.7 |

<sup>1</sup> AANHPI = Asian American, Native Hawaiian, Pacific Islander; AIAN = American Indian and Alaska Native; Multi = More than one race
<sup>2</sup> CKD = chronic kidney disease; CHF = congestive heart failure; COPD = chronic obstructive pulmonary disease; CVA = cerebrovascular accident; ESRD = end-stage renal disease ; TIA = transient ischemic attack

### Time trend in anticoagulant use

Trends over time in reports of anticoagulant use from 2007-2019 are shown in Figure 1. The use of warfarin started to decline in 2011 with the marketing of the first DOAC. Warfarin use continued to decrease while the use of DOACs increased over the remainder of our study. DOAC use was initially driven by dabigatran availability, but apixaban has been the predominant DOAC since 2016. By 2019, apixaban accounted for 66% of all DOAC use. Edoxaban use was not observed during our study period.

**Figure 1.**
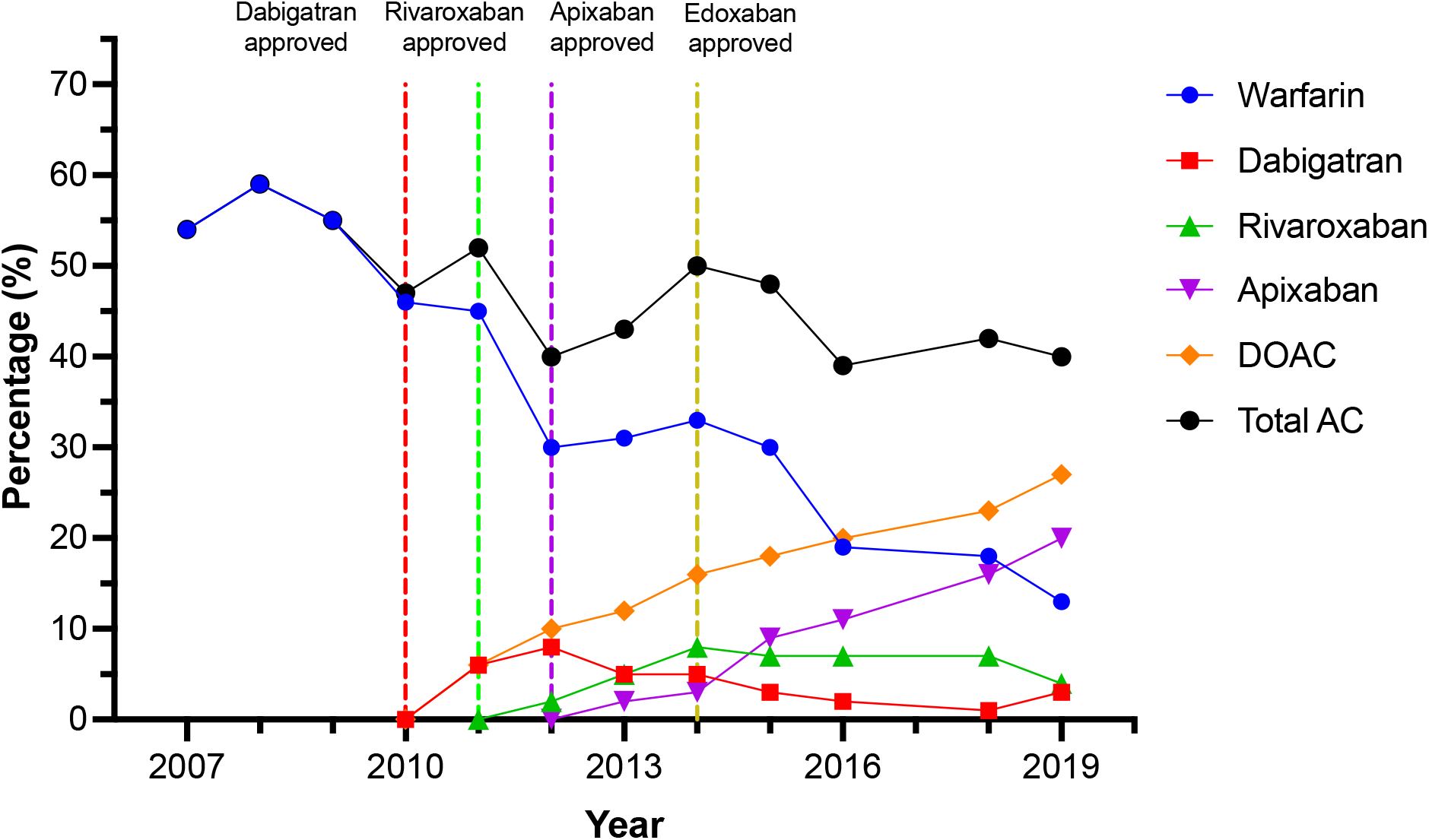
Percentage of US patients with atrial fibrillation and atrial flutter treated with direct oral anticoagulants, 2007-2019 AC= anticoagulant; DOAC = direct oral anticoagulant

### Factors associated with DOAC use

In multivariable analyses (Table 2), non-Hispanic Black patients appeared less likely to use a DOAC than non-Hispanic White patients (aPR: 0.74; 95% CI: 0.31-1.75), although the confidence interval was wide. Over the period 2011-2019, the prevalence of DOAC use for non-Hispanic Black patients declined over time relative to non-Hispanic White patients (aPR 0.75; 95% CI, 0.63-0.90). DOAC utilization among Hispanic patients (aPR 1.03; 95% CI, 0.84-1.25) or Asian American, Native Hawaiian, Pacific Islander, American Indian, or Alaska Native patients(aPR 0.97; 95%, 0.76-1.23) was similar to non-Hispanic White patients. Patients with Medicaid insurance were less likely to use a DOAC than those with private insurance (aPR 0.14; 95% CI, 0.04-0.46). The prevalence of DOAC utilization among those with Medicare insurance compared to those with private insurance was (aPR 0.85 95% CI, 0.450-1.43). Later years were associated with an increased likelihood of DOAC use (aPR 1.09; 95% CI, 1.00-1.19).

**Table 2.** DOAC Use and Its Association with Other Factors, 2011-2019.

| Characteristics | Adjusted<br>Prevalence Ratio (95%<br>CI) |
| --- | --- |
| Age | 1.00 (0.98-1.01) |
| Female | 0.83 (0.62-1.12) |
| Race<sup>*</sup> |  |
| White | 1 [Reference] |
| Black | 0.74 (0.31-1.75) |
| Hispanic | 1.13 (0.47-2.70) |
| AANHPI/AIAN/Multi | 1.39 (0.39-5.00) |
| Race x Year<sup>*</sup> |  |
| White | 1 [Reference] |
| Black | 0.75 (0.63-0.90) |
| Hispanic | 1.03 (0.84-1.25) |
| ANHPI/AIAN/Multi | 0.97 (0.76-1.23) |
| Medical Comorbidities<sup>†</sup> |  |
| CKD | 0.69 (0.39-1.23) |
| CHF | 1.34 (0.85-2.11) |
| COPD | 1.13 (0.52-2.52) |
| CVA/TIA | 0.65 (0.40-1.10) |
| Diabetes | 1.22 (0.72-2.06) |
| Hypertension | 1.43 (0.94-2.17) |
| Insurance |  |
| Private | 1 [Reference] |
| Medicare | 0.84 (0.50-1.42) |
| Medicaid | 0.14 (0.04-0.46) |
| Other | 0.96 (0.44-2.12) |
| Year | 1.09 (1.00-1.19) |

<sup>\*</sup> AANHPI = Asian American, Native Hawaiian, Pacific Islander; AIAN = American Indian and Alaska Native; Multi = More than one race
<sup>†</sup> CKD = chronic kidney disease; CHF = congestive heart failure; COPD = chronic obstructive pulmonary disease; CVA = cerebrovascular accident; ESRD = end-stage renal disease ; TIA = transient ischemic attack

### Sensitivity analysis

The sensitivity analysis, which included all the medications recorded (up to 30 medications) and all the diagnoses recorded (up to 5 diagnoses) in NAMCS, resulted in 63.1 million users of anticoagulants out of 121.5 million AF/AFL visits, compared to 46.5 million anticoagulant users out of 103.6 million visits in the primary analysis (Figure 2). The percentage of different races and ethnicities remain similar (non-Hispanic White 87.7 vs. 87.4%; non-Hispanic Black: 5.3 vs 5.5%; Hispanic 4.3 vs. 4.5%; AANHPI/AIAN/Multi: 2.6 vs. 2.6%) Once all of the medications and all of the diagnoses recorded between 2011-2019 were included in the sensitivity analysis, there is association between Non-Hispanic Black race/ethnicity and DOAC use over time relative to non-Hispanic White patients remains similar (aPR 0.91; 95% CI 0.74-1.12). Those with Medicaid insurance remain less likely to use DOACs compared to those with private insurance (aPR 0.16 95% CI 0.05-0.48). Non-Hispanic Black patients remain less likely to use a DOAC compared with non-Hispanic White patients (aPR: 0.48; 95% CI 0.20-1.17).

**Figure 2.**
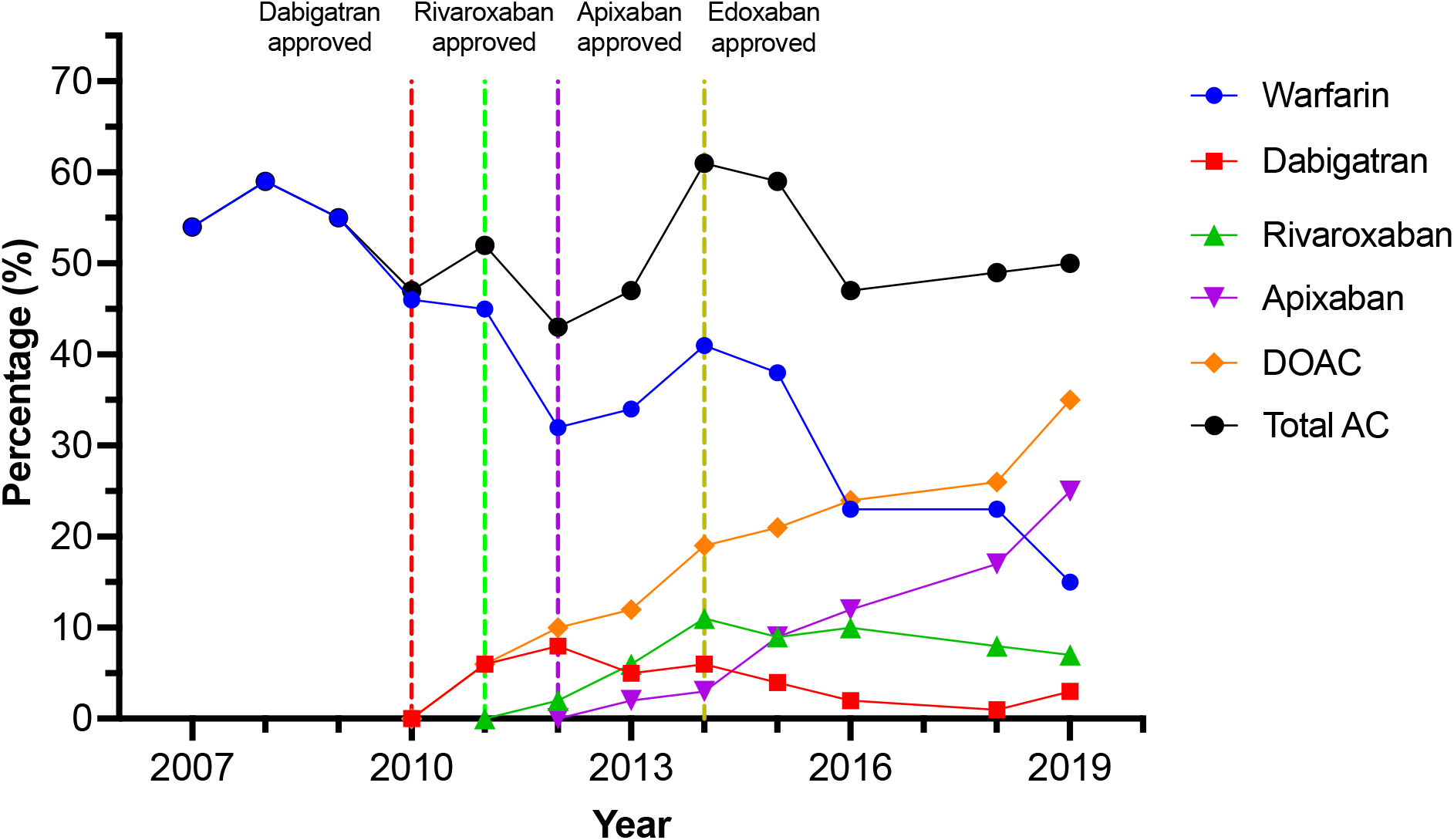
Sensitivity analysis of the percentage of US patients with atrial fibrillation and atrial flutter treated with direct oral anticoagulants, 2007-2019 AC= anticoagulant; DOAC = direct oral anticoagulant

## DISCUSSION

In our analysis of a nationally representative sample of ambulatory care visits, we observed significant racial and socioeconomic disparities in anticoagulant use among AF and AFL patients. Non-Hispanic Black patients and those with Medicaid insurance were less likely to use DOACs compared to their non-Hispanic White counterparts and those with private insurance.

Notably, the disparity in DOAC use for non-Hispanic Black patients worsened over time. This raises concerns about persistent inequalities in treatment access and outcomes. Concurrently, we observed a marked increase in the use of DOACs among patients diagnosed with AF or AFL from 2011 to 2019 that continued after the period covered by prior studies and four years beyond the most recent DOAC entered the market (edoxaban in 2015). Concurrently, the use of warfarin declined. This shift from warfarin to DOACs was predominantly influenced by a rise in apixaban use, which comprised two-thirds of all DOAC use in 2019. The prevalence of rivaroxaban and dabigatran use reached a plateau after apixaban entered the market and declined over the final years of our study period.

Our study has revealed socioeconomic and racial disparities in ambulatory care anticoagulant use among AF and AFL patients. We found that patients with Medicaid insurance were less likely to use a DOAC than patients with private insurance. Notably, non-Hispanic Black patients were using DOACs at a considerably lower rate than their non-Hispanic white counterparts, even after adjusting for the source of insurance coverage (a proxy for socioeconomic status) and other covariates. This disparity was not observed among Hispanic patients, who were similarly as likely as non-Hispanic white patients to use DOACs. These findings are consistent with prior research on imbalances of anticoagulant use in other population groups, underscoring a persistent disparity in recent years—a period that should reflect modern prescribing practices. For instance, the ORBIT-AFII study had shown that non-Hispanic Black patients had lower rates of DOAC utilization than non-Hispanic White patients, even after controlling for socioeconomic variables, including residential area, educational attainment, and insurance type.^7^ Furthermore, a study within the Veterans Health Administration System corroborated this racial disparity in AF patient treatment.^8^ Moreover, a study of a commercially insured population reported that non-Hispanic Black patients were less likely to receive DOAC therapy for incident venous thromboembolism.^9^ Socioeconomic factors, including lower household net worth, have also been found to be associated with a lower likelihood of being prescribed a DOAC among nonvalvular AF patients.^10^ A study using the Medical Expenditure Panel Survey on DOAC use during the early years of availability showed that increased DOAC use has been associated with higher out-of-pocket spending,^22^ further highlighting the socioeconomic barrier to accessing these medications. Our findings underscore the role of socioeconomic status and race in anticoagulation management for AF and AFL patients. While factors such as implicit bias and trust in medical care have been noted as contributors to care disparities, issues such as the high cost of DOACs and access to specialists who are more likely to prescribe DOACs play a crucial role as contributional factors in racial and ethnic disparities in contemporary care.^23–26^ Furthermore, chronic kidney disease, a condition that disproportionately affects non-Hispanic Blacks,^27,28^ might also influence prescribing patterns. |There is a critical need for research focused on identifying and addressing the root causes of these disparities and developing targeted strategies that ensure equitable access to specialized care, novel treatments, and emerging best practices for all. Furthermore, there is likely a role in medical education and continuing education of healthcare providers for shared decision-making, cost-conscious care, and implicit bias training to address these inequalities.^29–32^

Between 2016 and 2017, the prevalence of DOAC use surpassed that of warfarin among patients with AF or AFL, consistent with findings from prior research.^11,33^ The increase in DOAC use can be attributed to potential advantages in safety and effectiveness and to the convenience of DOAC use, as it obviates the need for regular anticoagulation monitoring and dietary restrictions. The initial uptake in DOAC adoption began with the FDA’s approval of dabigatran and rivaroxaban in 2010 and 2011, respectively. Subsequently, apixaban was approved in late 2012 and became the predominant contributor to the increase in DOAC use.

Several factors may influence the shift toward apixaban. Although data on the reversal of non-VKA oral anticoagulants are scarce, guidelines from leading societies recommend specific reversal agents for these medications in cases of life-threatening bleeding events such as intracranial hemorrhage. Specifically, the Neurocritical Care Society and the Society of Critical Care Medicine recommend using idarucizumab for dabigtran reversal and four-factor prothrombin complex concentrate (PCC) for oral direct factor Xa inhibitors reversal.^34^ While four-factor PCC has been commonly available in hospitals since its approval for VKA reversal in 2013, only 60% of the US hospitals providing emergency care have access to idarucizumab.^35^ The availability of anticoagulant reversal agents in the event of major bleeding can potentially affect prescriber and patient preferences.^36^ Additionally, concerns over the comprehensiveness of clinical trial data released for dabigatran and its safety profile may have further influenced prescribing patterns.^37–39^ Moreover, the preference for apixaban is likely bolstered by the ARISTOTLE (Apixaban for Reduction in Stroke and Other Thromboembolic Events in Atrial Fibrillation) trial outcomes,^40^ which showed its superiority over warfarin in reducing the risk of stroke, systemic embolism, major bleeding, and mortality in AF patients. In contrast, rivaroxaban was only found to be non-inferior to warfarin for these outcomes.^4^

Our analysis has several strengths and limitations. NAMCS is a nationally representative survey of non-federally employed, office-based physician visits. This strengthens the generalizability of our results when compared to single health system studies or studies with specific populations such as Medicare beneficiaries or patients in the Veterans Health Administration System. Nevertheless, our study has several limitations. First, the data collected in NAMCS is visit-level rather than patient-level, thus lacking granularity in certain areas, such as the patient’s complete past medical histories, zip codes, and household income levels. This does not allow us to control for unobserved covariates that may affect treatment selection, such as the degree of chronic kidney disease and risk scores (e.g., CHA_2_DS_2_-VASc, HAS-BLED…etc.) used to inform treatment decisions. Second, it is noted that the nonresponse rates for race and ethnicity for NAMCS have been reported to be between 20-30% from 2007 to 2019.^41^ NAMCS used model-based single imputation to address the missing race and ethnicity information. It is important to recognize that high percentages of nonresponse items may lead to biased estimates. Nonetheless, we believe that the association between DOAC usage and race found in our study aligns with existing literature on racial disparities in healthcare. However, they should be interpreted cautiously due to potential imputation bias. Third, the collection of visit-level data for NAMCS was disrupted and discontinued after 2019 due to the COVID-19 pandemic, precluding the extension of our analysis beyond 2019. Finally, a notable limitation arises from the evolution of NAMCS’s data collection methodology. Specifically, the number of medications and diagnoses documented by NAMCS expanded from 8 and 3 in 2007 to 30 and 5 by 2019, respectively. This affected the overall trend in anticoagulant usage when using only the first 8 versus all available fields. Our sensitivity analysis also revealed discrepancies when including a more comprehensive set of medications and diagnoses recorded in the NAMCS database, particularly affecting the statistical significance of associations observed in the primary analysis between non-Hispanic Black and DOAC use. These findings suggest that the broader medication and diagnosis data may introduce variables that dilute the observed associations.

Despite these differences, our primary analysis’s fundamental trends and disparities remain important. These initial results are robust and are consistent with previous research that has identified similar patterns in healthcare access and treatment decisions. Therefore, while the expanded dataset in the sensitivity analysis provides a valuable perspective that encourages a more nuanced understanding of the factors influencing anticoagulant use, the findings from our primary analysis continue to hold significant clinical and policy relevance. Future research utilizing NAMCS data should also account for this expanded medication and diagnoses recording framework in their analysis plan to ensure a comprehensive analysis of drug utilization trends.

## CONCLUSION

Amid a significant shift in the treatment for AF and AFL from warfarin to DOACs, non-Hispanic Black patients were far less likely than patients of other races and ethnicities to use these newer anticoagulant options. This observation underscores the necessity for further research to determine the underlying factors contributing to this inequality. Understanding these disparities is crucial for the development of focused strategies aimed at ensuring equitable access to novel treatments and emerging best practices for all.

### Sources of Funding

Dr. McManus is supported by grants from the National Institutes of Health (U54HL143541, R01HL141434, R33HL158541, and R01HL155343)

## Data Availability

All data referred to in this manuscript are available upon reasonable request All data used in population science research are derived from publicly available sources and can be accessed at https://www.cdc.gov/nchs/ahcd/datasets_documentation_related.htm. Specific data citations and links are provided within the manuscript for reference

